# PvGAP: Development of a globally-applicable, highly-multiplexed microhaplotype amplicon panel for *Plasmodium vivax*

**DOI:** 10.1101/2025.04.30.25326751

**Authors:** Alfred Hubbard, Edwin Solares, Lauren Bradley, Brook Jeang, Delenasaw Yewhalaw, Daniel Janies, Eugenia Lo, Guiyun Yan, Elizabeth Hemming-Schroeder

## Abstract

**Background:** *Plasmodium vivax* malaria research has yet to fully benefit from the advances in genomic surveillance that have revolutionized *P. falciparum* epidemiology. Closing this gap is critical because genomic tools are necessary to monitor the spread of drug resistance, classify infections as local or imported, and distinguish reinfection, recrudescence, and relapse. To achieve these objectives, microhaplotype marker panels that allow powerful genotyping of polyclonal infections are needed.

**Methods:** We designed a Globally-applicable Amplicon Panel for *P. vivax* (PvGAP), selecting targets based on both genetic diversity and genetic distance from each other to maximize discriminatory capability between geographic regions. We evaluated this panel with field samples from Ethiopia and *in silico* using whole genomes from the MalariaGEN Pv4 database.

**Results:** PvGAP has 80 high diversity targets suitable for population genomics and eight targets of specific epidemiological interest, such as putative markers of drug resistance. We demonstrate PvGAP achieves robust amplification with field data and that it provides competitive accuracy for relatedness inference in three disparate geographic regions.

**Conclusions:** PvGAP joins existing *P. vivax* panels as a cost effective and practical option for genomic epidemiology of this neglected disease. It will support drug resistance surveillance, discrimination of local and imported cases, and it may aid in separating reinfection, recrudescence, and relapse in therapeutic efficacy studies, all critical needs of National Malaria Control Programs.

## Background

*Plasmodium vivax* remains understudied compared to *P. falciparum*, despite the fact that it can cause severe and life-threatening malaria (Anstey et al., 2012) and is endemic in tropical regions throughout the world, including most of Africa (Twohig et al., 2019). Improving our understanding of *P. vivax* epidemiology is a key step to planning effective antimalarial interventions and ultimately eliminating malaria.

Population genomics is one powerful approach for enhancing our collective understanding of malaria epidemiology. Genomic data from malaria parasites can be used to assess transmission between geographic locations and can support modeling of relapse, recrudescence, and reinfection (Neafsey et al., 2021). However, the application of population genomics to malaria epidemiology is limited by technical constraints and costs. Classical biallelic SNP assays have low sensitivity to detect multiple parasite strains and parasite diversity within a host (Koepfli & Mueller, 2017). On the other hand, whole genome sequencing (WGS) data provides comprehensive coverage, but is costly to produce (Tessema et al., 2022) and store (Neafsey et al., 2021).

A cost-effective alternative with high sensitivity to detect minority clones is targeted deep sequencing of genetically-diverse amplicons (Koepfli & Mueller, 2017; Tessema et al., 2022). This approach is particularly effective when used to assess multiallelic microhaplotypes, meaning targets that contain two or more SNPs. Microhaplotype markers have been shown to provide higher power for relatedness inference than biallelic SNPs, particularly in the case of polyclonal infections (Tessema et al., 2022). The benefits of microhaplotype panels for understanding genetic relatedness and diversity have already been demonstrated in *P. falciparum* (LaVerriere et al., 2022; Tessema et al., 2022).

Several microhaplotype panels have been or soon will be published for *P. vivax*. Particularly noteworthy are the Kleinecke et al. (2025) and PvGTSeq (Manrique-Valverde et al., 2025) panels, which both contain putative markers of drug resistance and numerous high diversity targets (93 for Kleinecke et al. and 213 for PvGTSeq). A handful of other panels have been developed with smaller sets of microhaplotypes (Kattenberg et al., 2022, 2024; Popkin-Hall et al., 2024; Rosado et al., 2025). Given that biallelic markers, which comprise most of the Kattenberg et al. (2022), Kattenberg et al. (2024), and Popkin-Hall et al. (2024) panels, are fundamentally limited in the presence of polyclonal infections (Tessema et al., 2022) and the small size of the Rosado et al. (2025) panel (11 targets), these panels are likely to have less power for relatedness estimation than the PvGTSeq and Kleinecke et al. panels. They may retain sufficient power in certain settings and for other applications such as estimating the number of unique strains in an infection (i.e., the complexity of infection).

This paper describes the design and validation of a *Plasmodium vivax* Globally-applicable Amplicon Panel (PvGAP) that is similar in scope to the Kleinecke et al. (2025) and PvGTSeq (Manrique-Valverde et al., 2025) panels. PvGAP consists of 80 high-diversity microhaplotype markers, seven putative drug resistance markers, and a section of *pvdbp* (*P. vivax* Duffy binding protein). The associated library preparation protocol is relatively low cost and uses non-proprietary reagents. Therefore, PvGAP is a powerful and flexible addition to the *P. vivax* population genomics toolkit that should suit a variety of epidemiological applications.

## Methods

### Panel design

Whole genome sequence data from 198 *P. vivax* isolates from eight countries were downloaded from NCBI (Table 1). Reads were aligned to the PvP01 reference genome (Auburn et al., 2016) and variants were called using standard bioinformatic pipelines. Candidate loci were identified using a sliding-window method adapted from Tessema et al. (2022). Genomic windows were filtered to remove repetitive regions, indels, and loci lacking within-country polymorphism. Remaining windows were evaluated based on within-country nucleotide diversity (π) and population differentiation (FST) among countries. Windows with high diversity and/or strong genetic structuring were prioritized across chromosomes, yielding 278 candidate targets distributed across the genome. Detailed bioinformatic methods, filtering criteria, and marker selection procedures are provided in Supplemental Text 1.

**Table 1:**
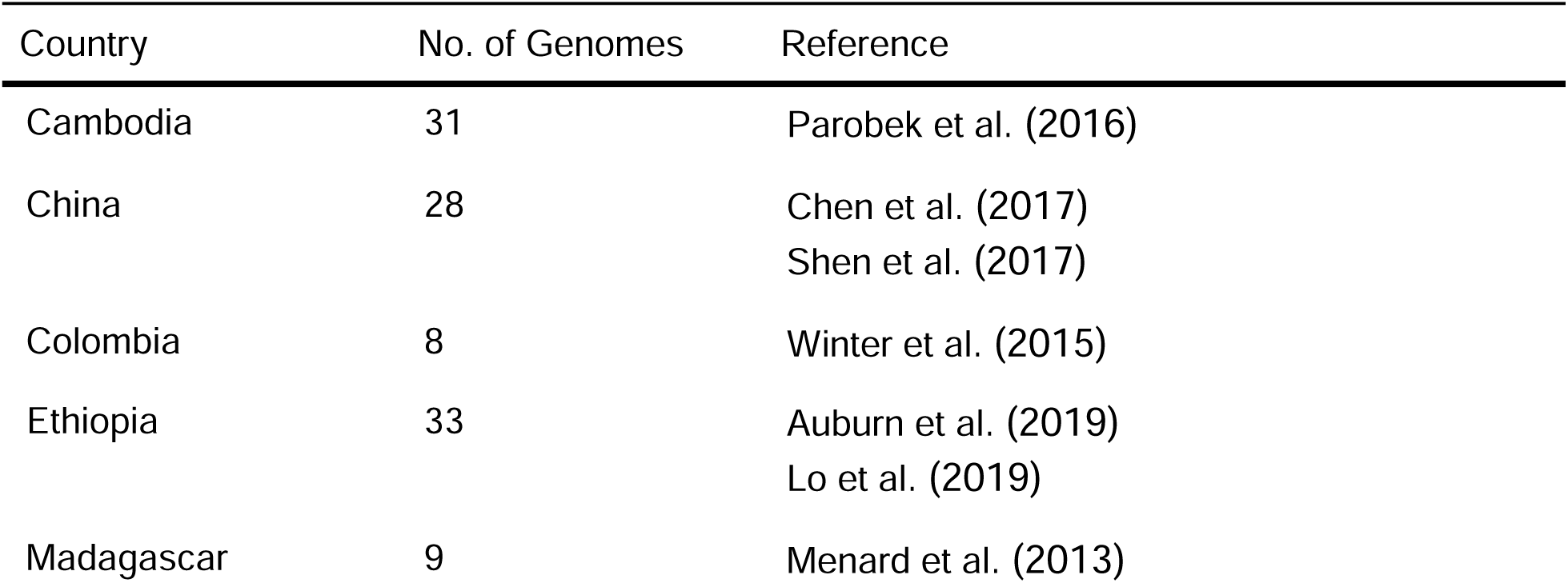
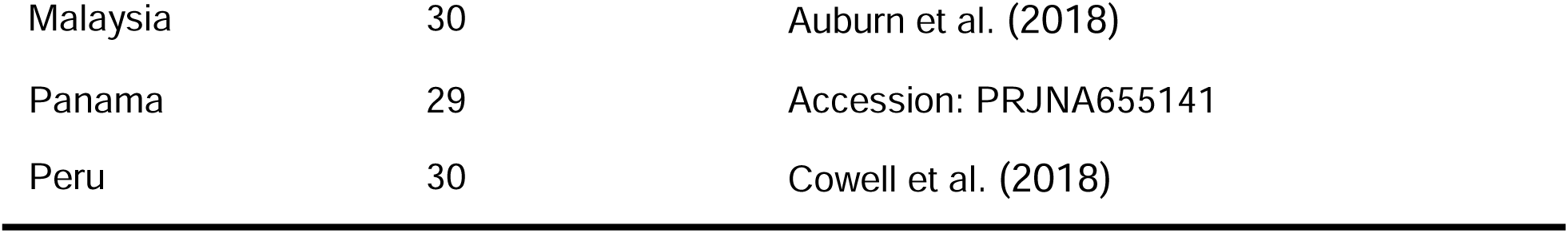
*P. vivax* genomes used in panel design.

| Country | No. of Genomes | Reference |
| --- | --- | --- |
| Cambodia | 31 | Parobek et al. (2016) |
| China | 28 | Chen et al. (2017)<br>Shen et al. (2017) |
| Colombia | 8 | Winter et al. (2015) |
| Ethiopia | 33 | Auburn et al. (2019)<br>Lo et al. (2019) |
| Madagascar | 9 | Menard et al. (2013) |
| Malaysia | 30 | Auburn et al. (2018) |
| Panama | 29 | Accession: PRJNA655141 |
| Peru | 30 | Cowell et al. (2018) |

These targets were submitted to GTseek LLC (Twin Falls, ID, USA) for multiplex primer design optimized to minimize primer crosstalk (Campbell et al., 2015). After initial sequencing tests, poorly performing targets were removed, resulting in a final set of 80 loci. Eight additional loci targeting *pvdbp* and putative drug resistance markers were added to the panel (Table 2), resulting in a final panel of 88 loci (Supplemental Data 1).

**Table 2:**
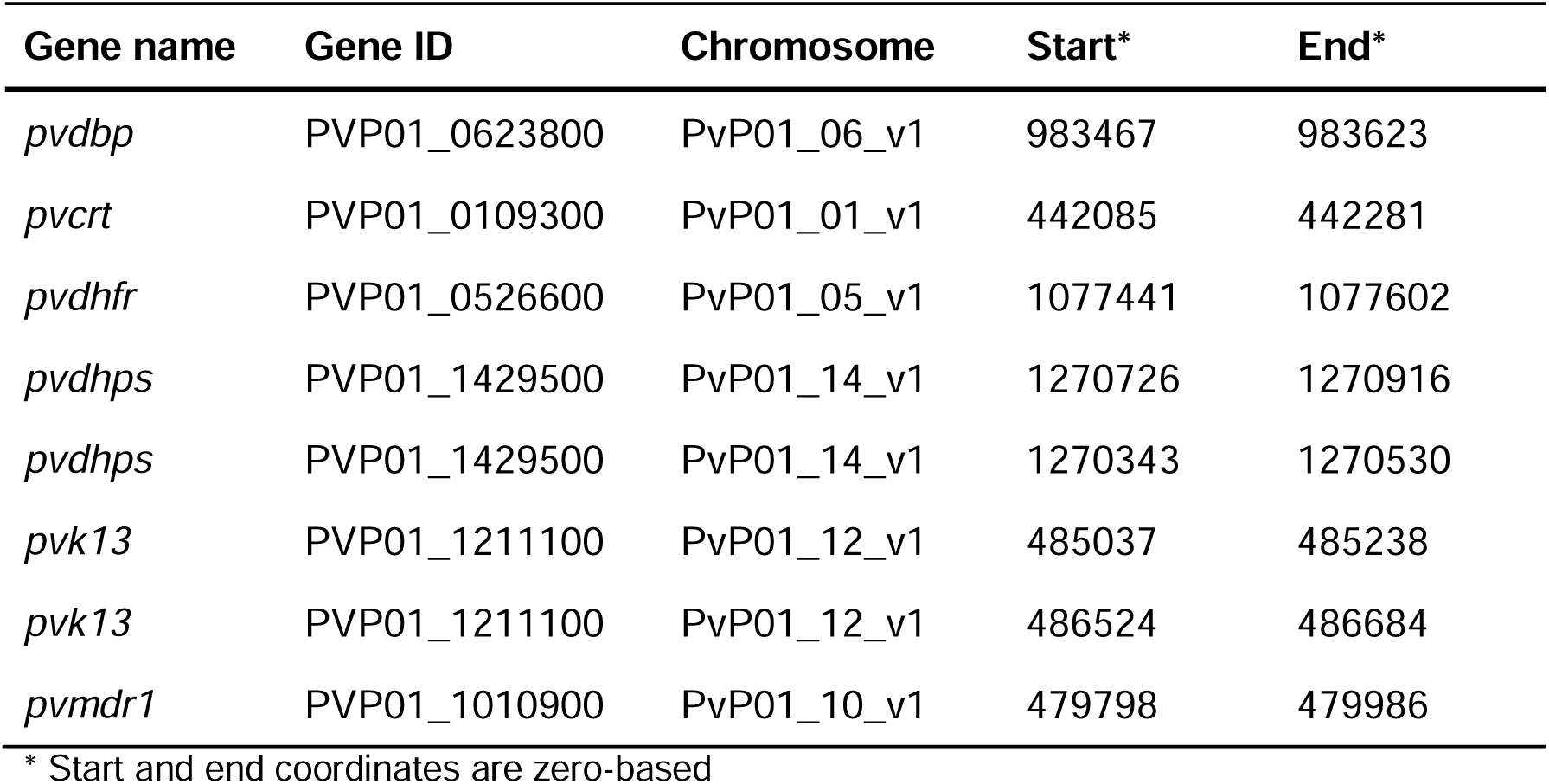
*P. vivax* genes of particular epidemiological interest.

### Panel evaluation with field samples

Two sets of field samples were used to evaluate sequencing performance of the panel. First, ten high-parasitemia samples (six dried blood spot (DBS) and four whole blood samples) were processed using multiple laboratory workflows to assess how different DNA extraction methods (Chelex/saponin versus spin column kit) and enrichment strategies (selective whole genome amplification or SWGA versus targeted pre-amplification) affected sequencing outcomes. A detailed description of these procedures is provided in Supplemental Text 2. Second, to assess how parasite density influences sequencing output, serial dilutions were generated from five high-parasitemia DBS samples to mimic realistic low-parasitemia infections. The procedures and results of this analysis are described in the main text below.

#### Sample collection

The samples used for the serial dilutions were gathered as part of a sub-Saharan Africa International Center for Excellence for Malaria Research (ICEMR) project (Githure et al., 2022; Yan et al., 2022). They were collected as DBS through both passive case detection and community cross-sectional surveys from two sites in Southwestern Ethiopia. Details on sample collection may be found in Getachew et al. (2023).

#### Library preparation and sequencing

DNA was extracted from DBS using the Monarch Spin gDNA extraction kit (New England Biolabs). Briefly, a 6 mm DBS punch was lysed and genomic DNA purified following the manufacturer’s protocol with minor modifications, with DNA eluted in 35 µL of TE buffer. Parasite DNA concentration was quantified by qPCR using primers and probes described by Lee et al. (2015) and a standard curve generated from serial dilutions of the *P. vivax* Sal I Sau3AI gDNA library (MRA-40, BEI Resources). Additional details of DNA extraction and quantification are provided in Supplemental Text 1.

Following quantification, samples were diluted to target parasite DNA concentrations of 1000, 500, 250, 100, 50, and 10 copies/µl. To generate serial dilutions that more closely mimic natural infections with low parasitemia (i.e., similar ratios of human to parasite gDNA), gDNA was extracted from uninfected human DBS following the same purification protocol and used as the diluent. Because a gDNA library was used as the quantification standard due to limited availability of parasite material, copy number estimates cannot be directly translated to parasitemia. However, assuming an approximate extraction efficiency of 30% and an input volume of 10 µL of whole blood per 6 mm DBS punch, a measured concentration of 10 parasite copies/µL of eluate would correspond to approximately 11.7 parasites/µL of whole blood. This estimate should be interpreted as an approximation given variability in DBS blood volume and extraction recovery.

The library preparation protocol is described in detail in Supplemental Text 3. In brief, it consists of SWGA, adapted from Oyola et al. (2016) and Cowell et al. (2017), to increase the density of parasite DNA; an adapted version of the GT-seq PCR protocol to affix primers (see Campbell et al. (2015) for the original technique and LaVerriere et al. (2022) for a previous application to *P. falciparum*); and application of Nate’s Plates kits (GTseek LLC) to affix dual indexing tags and normalize sequence quantity across samples. Various QC assays are performed after each step and on the final libraries, as described in Supplemental Text 3. Sequencing was performed on an Illumina MiSeq, using the Reagent Kit v2 500 cycles kit in paired-end mode with a 10% PhiX spike in. Samples were amplified and sequenced in triplicate.

#### Bioinformatics analysis

The malaria amplicon pipeline published alongside LaVerriere et al. (2022) was used to extract and filter microhaplotypes, using primer and reference files appropriate for our *P. vivax* panel (Supplemental Data 2). This pipeline uses the Divisive Amplicon Denoising Algorithm (DADA2; Callahan et al., 2016) as the core method for identifying and filtering amplicons. The pipeline then applies a handful of postprocessing steps to remove likely sequencing artifacts and chimeras.

Amplicons with fewer than five reads or a within-sample allele frequency (WSAF) below 1% were filtered out. Lin’s concordance correlation coefficient was computed among replicate pairs with the DescTools R package (Signorell, 2025).

### Panel evaluation with paneljudge and MalariaGEN data

#### Data preparation

To assess the utility of the panel for population genetics and genomic epidemiology, whole genome sequences from the MalariaGEN Pv4 project (MalariaGEN et al., 2022) were downloaded and virtual microhaplotypes were extracted. Variants that failed the QC process of the MalariaGEN authors (FILTER=“PASS” in the VCF) were excluded. Samples with an *F_WS_*below 0.95 were removed, as these are considered to be polyclonal (Auburn et al., 2012). Finally, samples from longitudinal studies and returning travelers were removed. From the remaining samples, three populations were selected based on sample size and representation of distinct regions: Cambodia and Vietnam in 2015 and 2016 (n=72); Brazil, Colombia, and Peru in 2013 and 2014 (n=43); and Ethiopia in 2013 (n=49).

The remaining variants were filtered to the genomic regions represented in PvGAP using bcftools view (Danecek et al., 2021). Haplotype sequences were obtained for each sample and locus with bcftools consensus (Danecek et al., 2021), using the PvP01 reference genome (Auburn et al., 2016). To enable comparison with other amplicon panels, the diversity targets of the Kleinecke et al. (2025) and PvGTSeq (Manrique-Valverde et al., 2025) panels were extracted with the same methods. Other *P. vivax* microhaplotype panels were excluded from this analysis because they either lack diverse microhaplotype targets (Kattenberg et al., 2022, 2024; Popkin-Hall et al., 2024) or are considerably smaller than the three panels included in the evaluation (Rosado et al., 2025).

#### paneljudge analysis

For each population, the allele frequencies of all panel targets were calculated with the Dcifer R package (Gerlovina et al., 2022). Cardinality (the number of unique alleles at a locus) and effective cardinality (the number of unique alleles adjusted for their frequency; see Taylor et al., 2019) were calculated from these frequencies with the paneljudge R package (https://github.com/aimeertaylor/paneljudge; v0.0.0.9). The paneljudge package was also used to simulate pairwise relatedness estimates for each panel under various true values of *r*: 0.01, 0.05, 0.1, 0.15, 0.2, 0.25, 0.5, 0.75, and 0.99. These simulations were performed with 100 sample pairs drawn from each population, 100 bootstrap replicates, and a switch-rate parameter value of 5.

### Multiplexing capacity

To estimate multiplexing capacity, we used locus-level read counts from the sequencing dataset generated in this study. Total on-target reads per sample and locus-specific read proportions were calculated, and relative sample efficiencies were derived to capture uneven sample representation in multiplexed libraries. We then used empirically parameterized Monte Carlo resampling to model expected locus coverage across Illumina sequencing kits with different read outputs. For each kit and candidate multiplex level, we assumed that 30% of reads would be lost (10% due to PhiX and 20% due to off-target amplification). We repeated the resampling procedure 100 times. Samples were considered to pass if at least 75% of loci achieved ≥10 reads, and recommended multiplexing levels were defined as the largest sample number for which ≥80% of samples were predicted to meet this criterion. Additional details are provided in Supplemental Text 5.

## Results

### Panel characteristics

Windows selected during panel design were chosen based on high nucleotide diversity and/or high F_ST_. Consistent with these criteria, all windows included in the final marker set exhibited high mean nucleotide diversity (median = 0.7552; IQR = 0.5555-0.9803; Figure 1A) and/or high mean F_ST_ (median = 0.4555, IQR = 0.31-0.5964; Figure 1B) in the genome set used for panel design.

**Figure 1:**
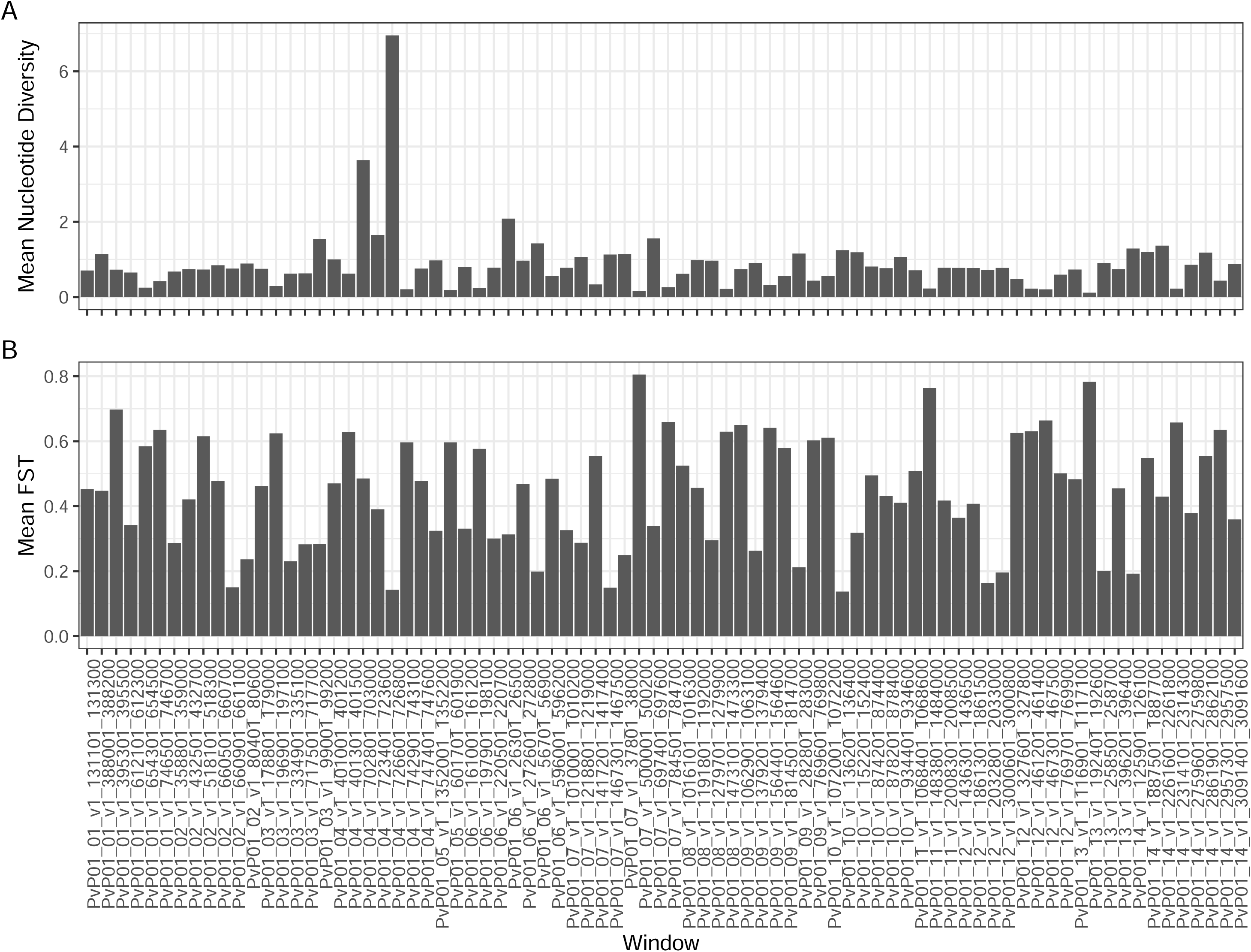
Mean nucleotide diversity **(A)** and mean F_ST_ **(B)** for the genome windows selected for inclusion in the final panel.

### Panel evaluation with field samples

Detailed results for the samples used for laboratory workflow optimization are described in Supplemental Text 2. In brief, both extraction methods performed similarly, and differences between enrichment strategies were modest. SWGA yielded a slightly higher proportion of on-target reads, whereas targeted pre-amplification resulted in slightly more even coverage across markers. SWGA and the Chelex/saponin method were selected for the final protocol.

In the serial dilution experiments, mean read counts across replicates (Supplemental Figure 1) indicate that overall amplification was satisfactory (median read depth per marker of 82; IQR = 22-391). A few loci exhibited lower amplification efficiency. Notably, these loci performed well in prior experiments (Supplemental Text 2), suggesting that this reduced performance is unlikely to represent intrinsic issues with the primers. Despite the reduced sequencing output at these loci, the proportion of targets with a minimum of 10 reads remained above 75% among all dilutions (Figure 2), suggesting the SWGA protocol enables reliable amplification in low parasite copy number samples. Moreover, off-target reads were low, with a median of 79.75% on-target reads per sample (IQR = 71.68-80.85%).

**Figure 2:**
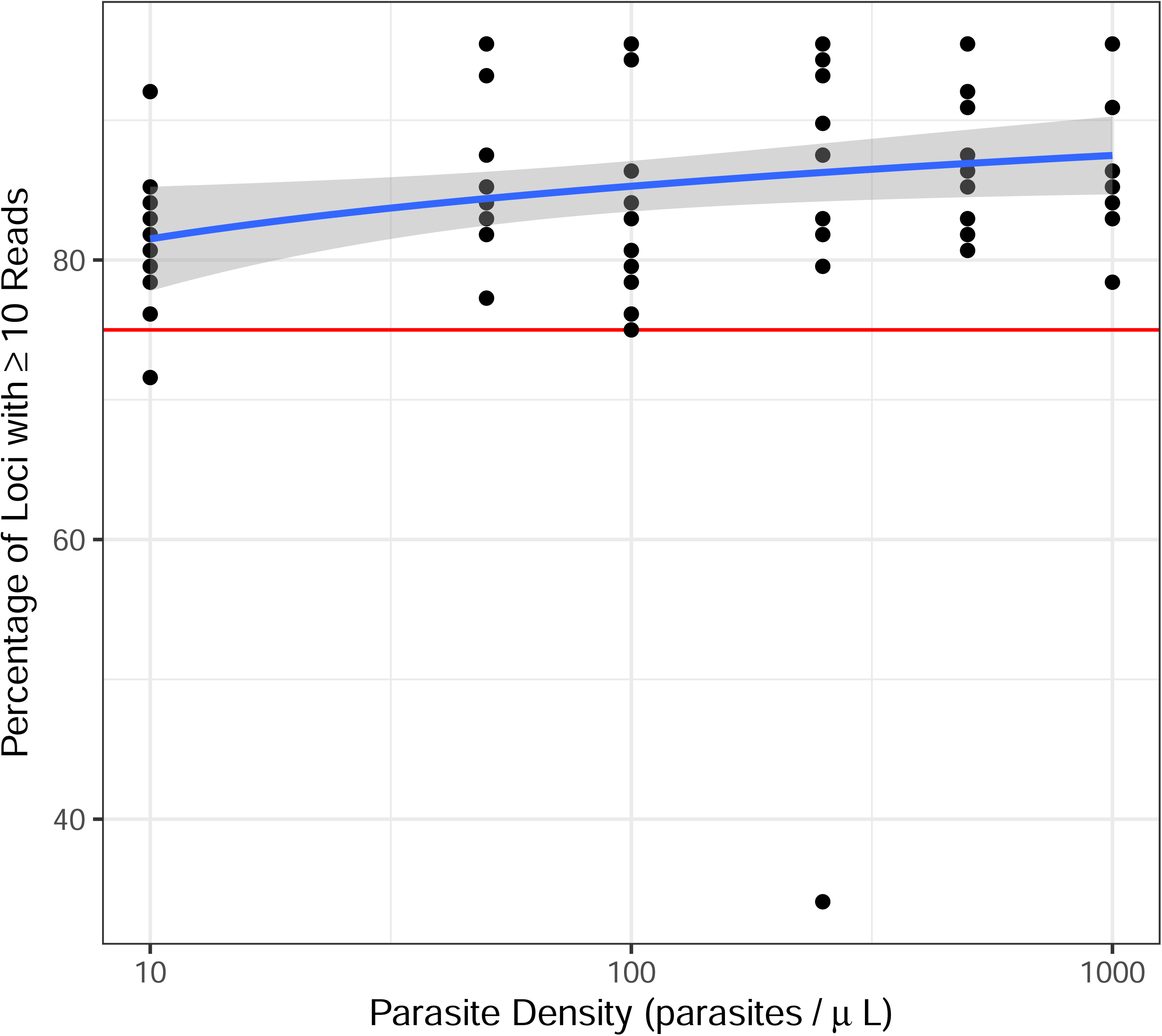
Scatterplot showing the relationship between parasite copy number and the percentage of loci with 10 or more reads (indicating successful amplification) for each replicate of the serial dilutions. The *x*-axis is log_10_ scaled. The blue line is a simple linear regression, with shaded areas showing 95% confidence intervals. The horizontal red line indicates 75% successful amplification, a common threshold for resequencing a sample.

Detection of minority alleles across replicates was inconsistent, though not strongly affected by parasite copy number (Supplemental Figure 2). Concordance among replicate pairs ranged widely, from nearly 0 to 1 when computed only on alleles present in both replicates of the pair (Supplemental Figure 3). Notably, concordance varied substantially among samples, possibly reflecting the presence of minor parasite strains at low densities or variation introduced during dilution preparation. When alleles only found in one replicate were included, concordance ranged from 0 to 0.6.

### Panel evaluation with paneljudge and MalariaGEN data

Comparison of the PvGAP panel with the panels from Kleinecke et al. and PvGTseq revealed that cardinalities (i.e., the number of unique alleles at each target) followed a similar pattern, with most markers showing a cardinality below 10 in most populations (Figure 3). Larger panels tended to have more high cardinality markers, though; PvGTSeq contains several markers with a cardinality above 20. In all panels, target effective cardinalities were fairly close to their theoretical maximum, the cardinality, suggesting the targets have sufficient allelic evenness to be informative for relatedness analyses (Figure 3).

**Figure 3:**
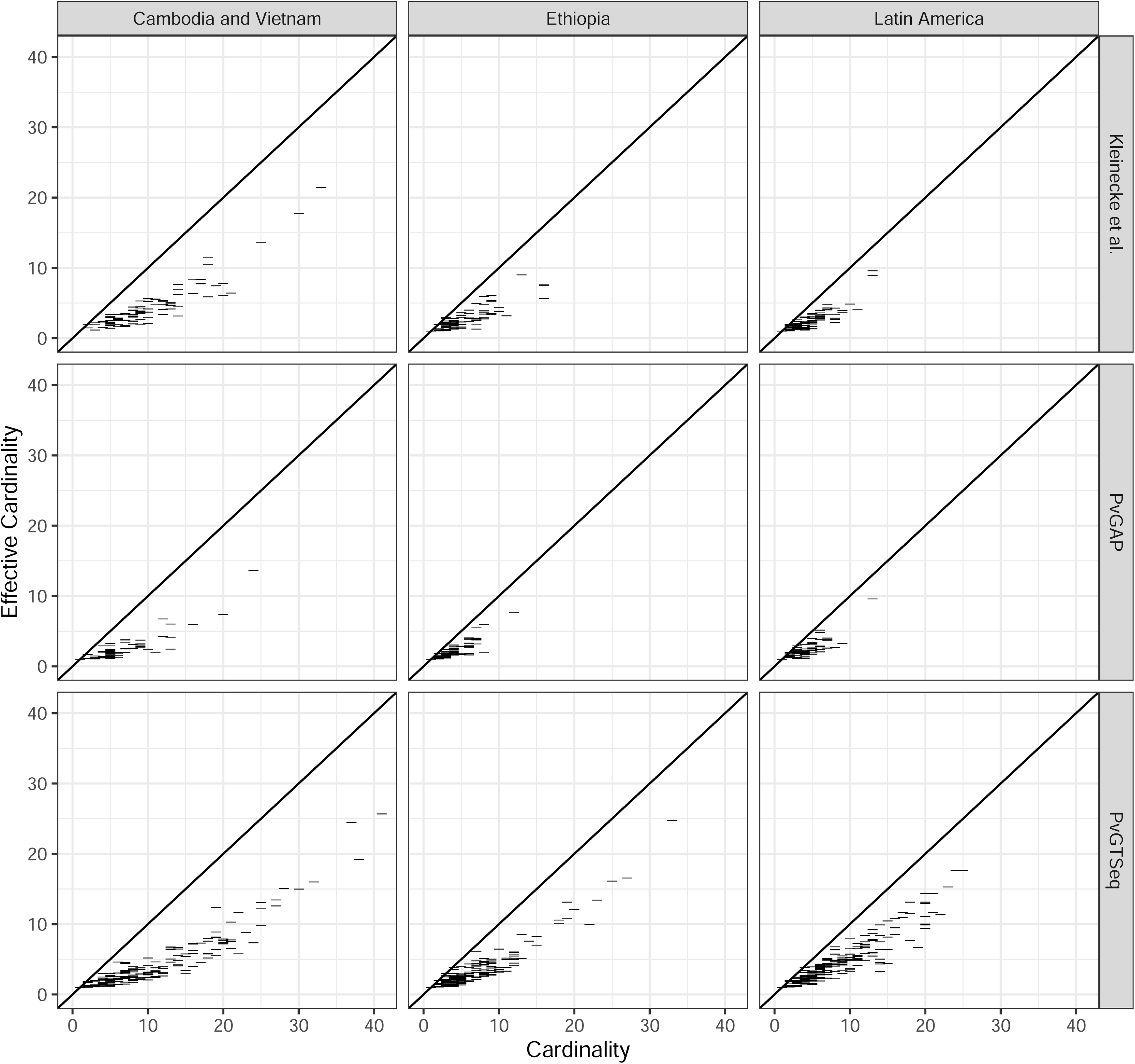
Cardinality (the number of unique alleles) versus effective cardinality (the number of unique alleles adjusted for allele frequency) for each marker (horizontal ticks indicate markers), based on the MalariaGEN data. The diagonal line shows *y=x*, which is the theoretical maximum of effective cardinality. The facets show the markers separately for each panel and population.

The paneljudge simulations indicate all three panels have substantial power for relatedness estimation. The RMSEs of 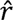 were 0.0625 to 0.125 for data-generating *r* values between 0.15 and 0.75 and lower for data-generating *r* values near zero or one (Figure 4). Similarly, the 95% confidence interval widths of these 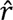 estimates tend to be wider (0.2-0.6) for intermediate data-generating *r* values and narrower near zero and one (Supplemental Figure 4). As expected from the number of diversity markers, PvGTSeq consistently yielded the lowest RMSE, followed by the Kleinecke et al. panel, followed by PvGAP. The differences are larger for intermediate data-generating *r* values and negligible for a data-generating *r* value of 0.99, indicating all panels are equally equipped to identify clonal pairs. PvGTSeq and PvGAP exhibit little difference in performance between geographic regions; the Kleinecke et al. panel exhibits slightly better performance in Southeast Asia than Ethiopia or Latin America. Notably, in all instances, for all panels, the 95% confidence interval widths are substantially less than one.

**Figure 4:**
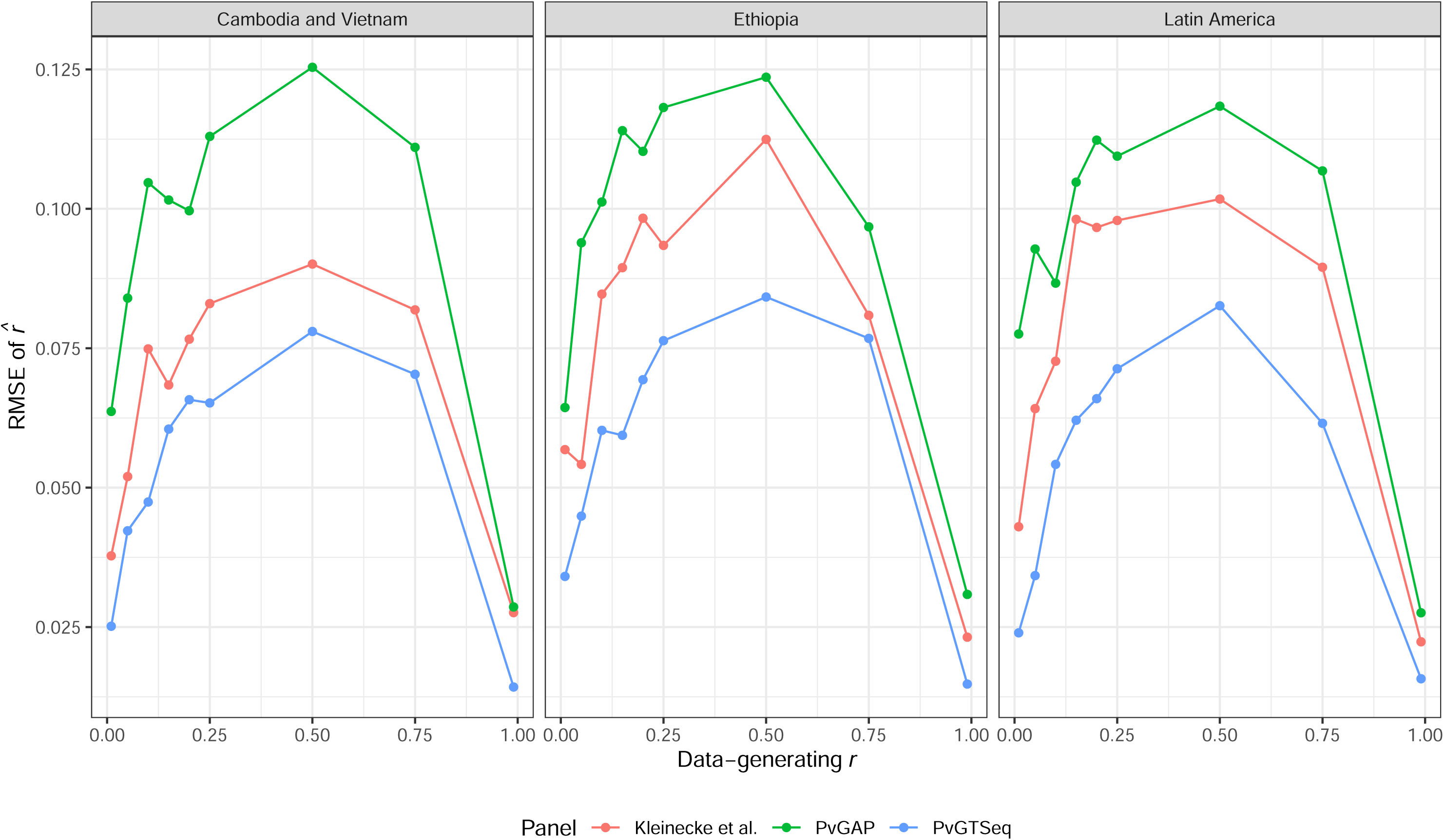
The RMSE of 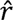 for each data-generating *r* value, as simulated by paneljudge. Facets distinguish results for different populations. Line and point colors indicate different marker panels.

### Multiplexing capacity and sequencing cost

Based on sample-to-sample unevenness, on-target read percentages, and locus-specific read counts observed in our dataset, we simulated multiplexing performance across Illumina sequencing kits to estimate feasible multiplexing levels and associated costs. For the Miseq v2 platform, the recommended multiplexing level is 136 samples per run, corresponding to an estimated sequencing cost of 13.24 USD per sample and a combined library preparation and sequencing cost of 28.24 USD per sample. Additional details and results for other sequencing kits are described in Supplemental Text 5.

## Discussion

PvGAP represents a cost-effective approach to targeted sequencing of *P. vivax* microhaplotypes for genomic epidemiology applications in varied geographic locations. The panel consists of 88 loci, including 80 selected for population genetic and relationship inference and eight within genes of epidemiological interest (e.g., potential drug resistance/tolerance markers). Our assay consistently yields reliable sequencing depth and breadth of coverage, even in the case of samples with low parasite copy numbers (10 parasites/μL), and does not rely on proprietary reagents. *In silico* evaluation with MalariaGEN data and paneljudge demonstrates that PvGAP contains sufficient marker diversity for relatedness estimation in a variety of geographic regions, albeit with lower accuracy than larger panels.

PvGAP displayed strong sequencing performance in terms of proportion of successfully amplified loci (all serial dilutions achieved >75% loci with ≥10× coverage) and percentage of on-target reads (median 79.75%). The former metric compares favorably to similar assays (Kleinecke et al. and PvGTseq). Multiplexing levels and on-target read percentages were not consistently reported by Kleinecke et al. (2025) and Manrique-Valverde et al. (2025), limiting direct comparison of sequencing efficiency and scalability of assays, particularly for DBS-derived samples. Regarding reproducibility across technical replicates, major allele detection was highly reproducible, but concordance of WSAFs varied among samples. This result may stem from stochastic detection of low-frequency minority clones, but this cannot be confirmed as these are field samples with unknown true WSAFs. Future benchmarking with synthetic mixtures could clarify interpretation of WSAF estimates and the need for further methodological refinements, if any.

In terms of relatedness estimation, the paneljudge analysis showed good performance of PvGAP, albeit slightly worse than similar, larger panels. All three panels evaluated (PvGAP, PvGTSeq, and Kleinecke et al) perform well when the true relatedness is high or low and somewhat worse when the true relatedness is intermediate (between 0.15 and 0.75). Although PvGTSeq consistently yielded the lowest RMSE, and PvGAP the highest RMSE (ranging from 0.028 to 0.125 depending on relatedness level and region), the importance of reducing RMSE should be evaluated in light of study objectives, budgetary considerations, and the biological limits inherent to malaria transmission inference. At present, the degree of precision required to reliably distinguish specific transmission events remains incompletely defined and likely varies by epidemiological setting and sampling design. Further empirical and simulation-based work is needed to determine the added value of sequencing larger, more costly panels that enable more accurate relatedness estimates.

Regarding cost, PvGAP is an affordable and flexible option. Library preparation for PvGAP is approximately 13 USD per sample (excluding primer costs; see below), compared to roughly 10-20 USD (converted from AUD) for the Kleinecke et al. (2025) assay, depending on whether the PCR reaction volume is halved. Once sequencing is included, we estimate a combined cost of approximately 28 USD per sample for PvGAP in a Miseq v2 run. Manrique-Valverde et al. (2025) report an estimated cost of approximately 39 USD per sample for library preparation and sequencing for PvGTSeq. Primer synthesis constitutes an upfront cost that scales with the number of targets. We estimate each locus requires approximately 24 USD in primer synthesis. Assuming the same cost for PvGTSeq, initial investment increases by several thousand USD. Finally, like the PvGTseq panel, the PvGAP primers have been designed to minimize cross-talk during multiplexing and library preparation by relying on standard amplicon-based methods rather than vendor-specific capture chemistries. In contrast, the Kleinecke assay relies on IDT’s proprietary rhAmpSeq methodology (Kleinecke et al., 2025), which may influence cost structure, supply-chain flexibility, and implementation considerations.

PvGAP is a moderately-sized panel for *P. vivax* epidemiology that provides a practical balance of feasibility and resolution for relatedness inference. In comparison to similar, previously-published panels, PvGAP offers competitive cost, slightly worse accuracy for relatedness estimation due to fewer markers, and a flexible laboratory protocol that can be adapted to a variety of settings. By substantially expanding the array of available panels, PvGAP helps ensure that the appropriate tool will be available for every application of genomic epidemiology to *vivax* malaria.

## Author Contributions

AH, EHS, and GY conceived of the study. DY and GY oversaw sample collection. EHS and ES designed the amplicon panel. LB, BJ, and EHS performed the laboratory work. AH performed data analyses and drafted the manuscript. DJ, EL, and EHS contributed to the interpretation of results. All authors provided critical feedback and helped shape the research, analysis, and manuscript.

## Acknowledgements

We thank the study participants and the field team of the International Centers of Excellence for Malaria Research (ICEMR) program for their involvement in this study.

## Funding Sources

This work was supported by NIH grants U19AI129326, F32AI147460, R21AI190606, and R01AI162947 and the Lucille P. and Edward C. Giles Dissertation-Year Fellowship.

## Conflicts of Interest

The authors declare no conflicts of interest.

## Data Availability Statement

The code necessary to generate all figures from the raw data is available on GitHub (https://github.com/a-hubbard/vivax_microhap) and Zenodo (will upload and include link once accepted). The raw sequence reads from the serial dilution experiment are available on SRA under Accession # PRJNA1435528. The references for the genomes used in panel design are described in Table 1, and the genomes used in panel evaluation were obtained from https://www.malariagen.net/resource/30/.

## Notes

### Competing Interest Statement

The authors have declared no competing interest.

### Author Declarations

The IRBs of University of California at Irvine; Case Western Reserve University, Cleveland, OH; and the Institute of Health of Jimma University, Ethiopia gave ethical approval for this work.

### Summary of Updates

Two major changes have been made to the manuscript: 1) the previous field data has been replaced with a serial dilution experiment to explore amplification in low parasite copy number samples; and 2) the MalariaGEN analysis has been reworked to use the paneljudge R package to assess accuracy for genetic relatedness estimation. Minor changes to organization and writing have also been made.

