## Supplemental Figures for "PvGAP: Development of a globally-applicable, highly-multiplexed microhaplotype amplicon panel for *Plasmodium vivax*"


Alfred Hubbard Edwin Solares Lauren Bradley Brook Jeang
Delenasaw Yewhalaw Daniel Janies Eugenia Lo Guiyun Yan
Elizabeth Hemming-Schroeder


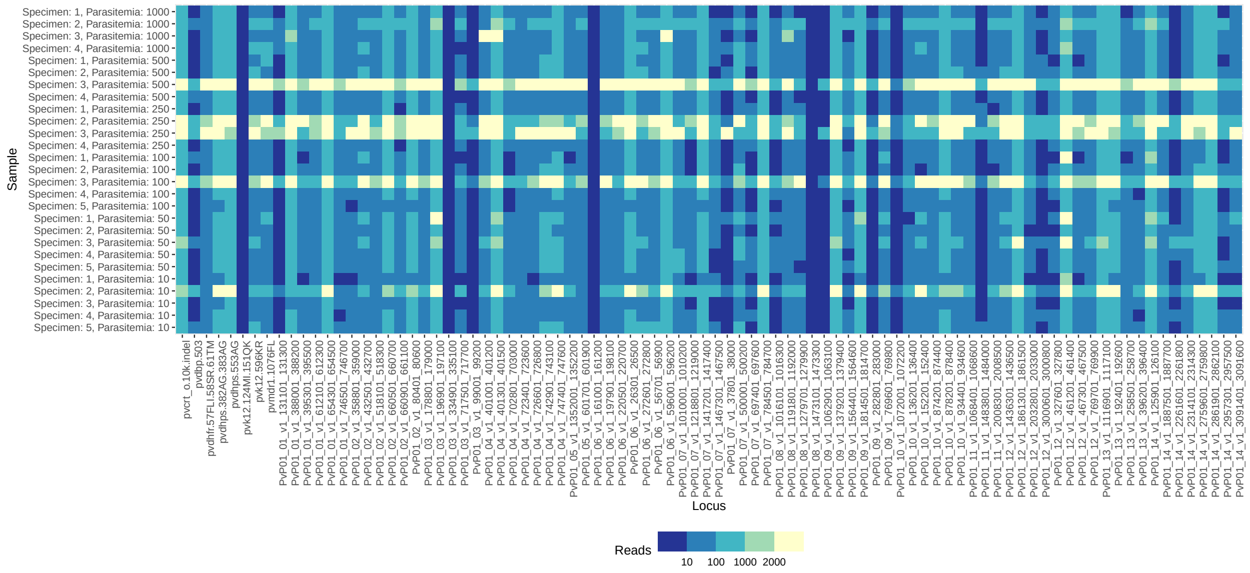


**Supplemental Figure 1:** Mean read counts across replicates for the serial dilutions, visualized separately for each locus. Values in the color legend indicate break points between bins. The $y$-axis tick labels indicate the specimen ID and parasite copy number of each dilution, with the latter in units of parasite copies/$\mu$L.

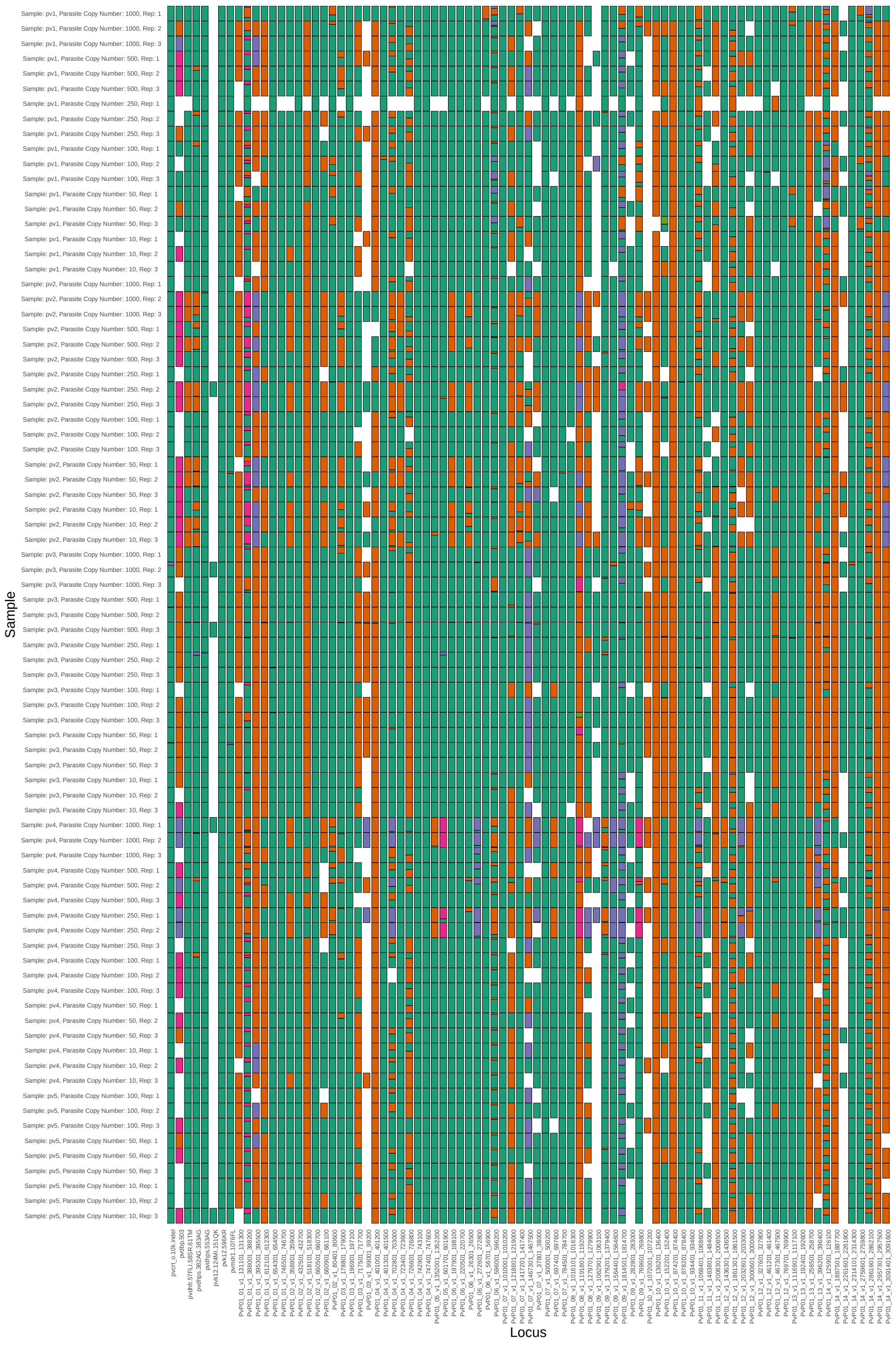


**Supplemental Figure 2:** Within-sample allele frequencies of the genotypes observed from each replicate. Rows correspond to replicates for each specimen and parasite copy number combination (in units of parasite copies/$\mu$L). Columns correspond to loci. Colored rectangles show the within-sample allele frequency of each allele for that replicate and locus. Within each column, the same colors correspond to the same alleles for that locus. Across columns, the color palette is repeated.

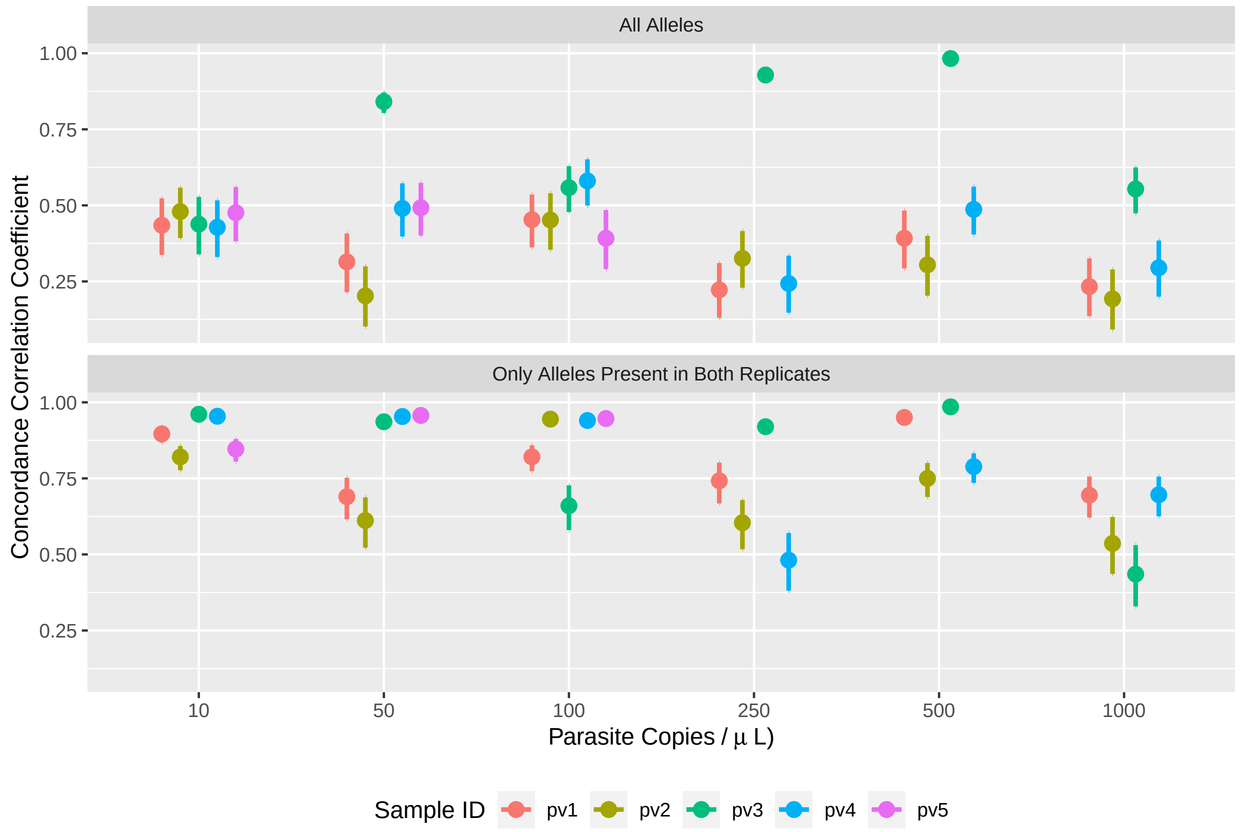


**Supplemental Figure 3:** Lin's concordance correlation coefficient between replicate pairs for each parasite copy number. The bars around each point indicate the 95% confidence interval of the estimate. Different colors distinguish different samples. Note that not all samples were evaluated at all parasite copy numbers. The top facet shows the concordance for all alleles, whereas the bottom shows the concordance calculated only on alleles that are present in both replicates of each pair.

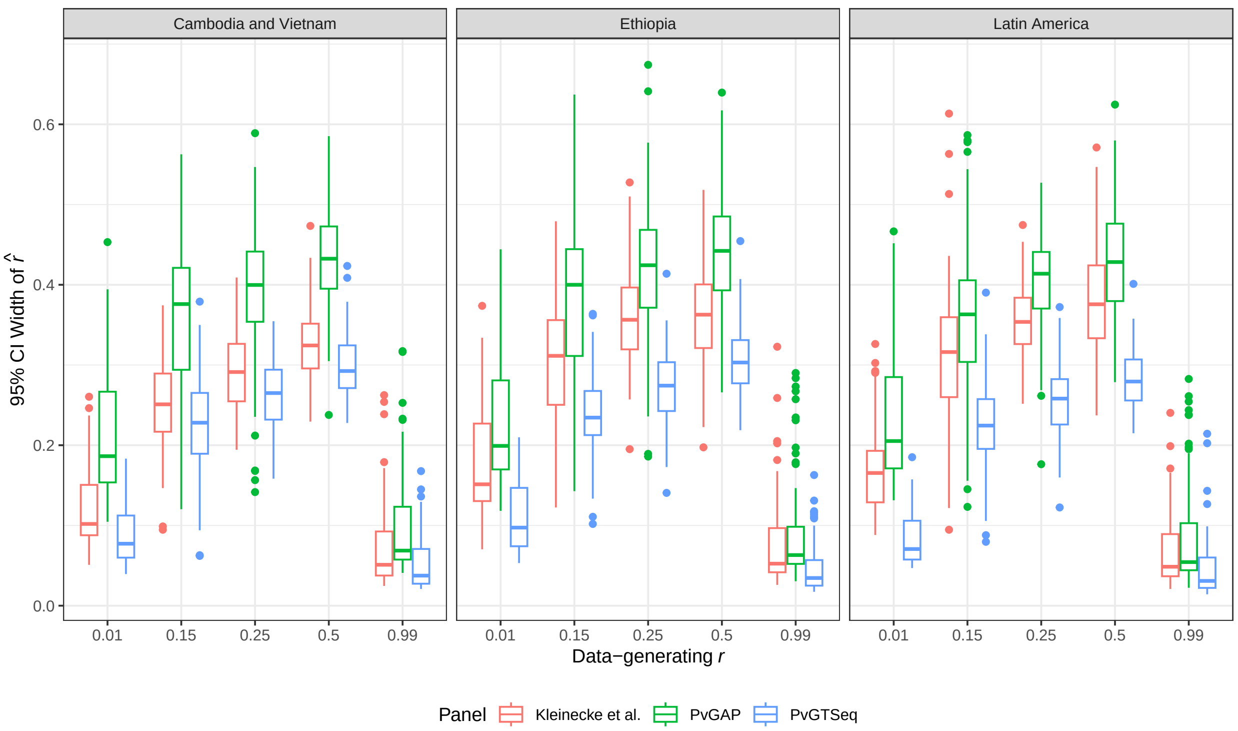


**Supplemental Figure 4:** Distribution of the 95% confidence interval widths of $\hat{r}$ for each data-generating $r$ value, as simulated by paneljudge. Facets separate results for different populations, and boxplot color identifies the marker panel. For clarity, only data-generating $r$ values of 0.01, 0.15, 0.25, 0.5, and 0.99 are included. The whiskers are Tukey-style and extend to a maximum of 1.5 * IQR.
