## Supplemental Text 1 for "PvGAP: Development of a globally-applicable, highly-multiplexed microhaplotype amplicon panel for *Plasmodium vivax*"

### Supplemental Text 1: Supplemental Methods

#### Panel design

Downloaded whole genome sequences were aligned with bwa (Li & Durbin, 2009) and SAMtools (Danecek et al., 2021). Alignments were removed using BCFTools (Danecek et al., 2021) if they had multiple mappings, mappings across chromosomes, mean coverage greater than 4x, or quality values less than 20. SNPs and indels were called using GATK (Van der Auwera & O’Connor, 2020), Picard-tools (*Picard Toolkit*, 2019), and VCF-tools (Danecek et al., 2011). Specifically, the HaplotypeCaller and GenotypeGVCFs algorithms from GATK were used.

To identify candidate markers, we used a sliding window method adapted from Tessema et al. (2022). We divided the PvP01 reference genome (Auburn et al., 2016) into 200bp windows every 100bp using the sliding.window.transform function in the PopGenome R package (Pfeifer et al., 2014). This yielded 242,135 windows, which were then passed through several filters. First, tandem repeats were identified using Tandem Repeats Finder (Benson, 1999). Windows that contained tandem repeats greater than 40 bp, dinucleotide repeats greater than 8 bp, homopolymer repeats greater than 8 bp, or trinucleotide repeats greater than 12 bp were removed. Second, windows containing any insertion or deletion from variant calling were removed. Third, within-country nucleotide diversity (π) was calculated using PopGenome, and windows that were monomorphic (π = 0) in isolates from greater than 25% of the countries (i.e., within more than two of the eight countries) were excluded, as these windows would not be informative for fine-scale genomic analyses in those countries. After filtering, 2,498 candidate windows remained.

We next evaluated and compared candidate windows based on polymorphism and genetic structuring. Mean within-country nucleotide diversity (π) and average fixation index (F_ST_) were calculated for each country against all other individuals. Both values were calculated in PopGenome using the F_ST.stats function. Next, for each chromosome, the 10 windows with the highest π, the window with the highest mean F_ST_, and the window with the highest π that was also in the highest 8% of F_ST_ values were selected. After that, the windows were ranked by π and F_ST_ across all chromosomes, and the 72 windows with the highest π, the 28 windows with the highest F_ST_, and the 10 windows with the highest π that were also in the highest 8% of F_ST_ values were selected. At this point, windows were removed from this set if there was insufficient availability of conserved regions outside of the target window for primer design and replaced with the window having the next highest value.

The final set of candidate windows which were selected for primer design consisted of 278 targets, with 16 to 25 targets on each chromosome. The minimum π for selected targets was 0.0024 and for F_ST_ the minimum was 0.50. These minimum values were among the top 55% and 93% of values, respectively, of the original 2,498 candidate windows.

These 278 targets were submitted to GTseek LLC (Twin Falls, ID; [https://gtseek.com](https://gtseek.com/)) to design primers that generate minimal crosstalk during multiplexed PCR reactions (Campbell et al., 2015). Primers were successfully designed for 179 of the 278 targets. After small test sequencing runs, we further removed primers for targets that did not amplify consistently, yielding a reduced set of 80 targets.

In addition to the targets selected for their π and/or F_ST_ values, eight additional loci of interest were added to the panel, bringing the final panel size to 88. These additional loci target *pvdbp* and putative markers of drug resistance. Primers for these targets were also designed by GTSeek LLC to minimize crosstalk among primers during amplification.

#### Library preparation and sequencing

DNA was extracted from DBS samples using the Monarch Spin gDNA extraction kit (New England Biolabs). A standard 6 mm DBS punch was incubated in 180 µL PBS at 85°C for 10 min. Samples were then incubated at 56°C following addition of 10 µL Proteinase K (1 mg/mL), 3 µL RNase A, and Blood Lysis Buffer. Genomic DNA was subsequently purified according to the manufacturer’s protocol and eluted in 35 µL TE buffer.

DNA concentration was quantified by qPCR using a standard curve generated from serial dilutions of the *P. vivax* Sal I Sau3AI gDNA library (MRA-40, BEI Resources) ranging from 10,000 to 1 copy per µL. Each reaction had a total volume of 10 µL and contained 5 µL Luna Universal Probe qPCR 2× Master Mix (New England Biolabs), primers and probe at 10 µM each, and 2 µL template DNA. Primers and probes followed Lee et al. (2015). Cycling conditions followed the manufacturer’s recommendations for the Luna Universal Probe qPCR Master Mix and were performed on a qTOWER 384 Real-Time PCR System (Analytik Jena).
