## Supplemental Text 2 for "PvGAP: Development of a globally-applicable, highly-multiplexed microhaplotype amplicon panel for *Plasmodium vivax*"

Supplemental Text 2: Small Batch Testing to Optimize Protocol

Alfred Hubbard Edwin Solares Lauren Bradley Brook Jeang
Delenasaw Yewhalaw Daniel Janies Eugenia Lo Guiyun Yan
Elizabeth Hemming-Schroeder

This document describes a series of small test runs that were performed with a handful of field samples to optimize the PvGAP protocol for DNA extraction and library preparation.

### 1 Methods

#### 1.1 Sample collection

To evaluate the panel in terms of sequencing yield and ability to obtain haplotypes, we used six samples previously collected as part of an ongoing Sub-Saharan Africa International Center for Excellence for Malaria Research (ICEMR) project. Some of these samples were obtained with passive case detection and some came from mass blood surveys. The samples were collected from three field sites in the Oromo region of Southwest Ethiopia; the Arjo-Didessa sugarcane farm, the Gambella region, and Jimma town.

In addition, four whole blood samples collected from Agaro Health Center, which is near Jimma, Ethiopia, were used to evaluate panel performance with DNA extracts derived from whole blood.

#### 1.2 Library preparation

For DBS samples, two different extraction methods were tested: the Qiagen QIAamp DNA Investigator Kit (from now on referred to as the "kit") and the Chelex/saponin method (Bereczky et al. 2005). For DBS samples, two different extraction methods were tested: the Qiagen QIAamp DNA Investigator Kit (referred to as the "kit") and the Chelex/saponin method (Bereczky et al. 2005). For the Chelex/saponin method, DNA extraction followed the protocol described by Bereczky et al. (Bereczky et al. 2005), except that the final elution volume was 200 µL. For the kit-based extraction method, we followed the manufacturer's protocol for "Isolation of Total DNA from FTA and Guthrie Cards," except that the final elution volume was 200 µL.

In addition, for all samples, three different enrichment strategies were tested: no enrichment, a targeted nested PCR prior to the GT-seq PCR (from here forward referred to as targeted pre-amplification), and selective whole genome amplification (SWGA) prior to the first PCR in the GT-seq protocol.

When SWGA was used, we performed the reaction and bead cleanup according to the established protocol (Cowell et al. 2017). When targeted pre-amplification was performed, nested reaction mixtures and amplification were conducted following the same conditions as PCR 1 of the GT-seq protocol.

Following enrichment, the protocol consists of an initial PCR with primers for amplicon targets containing adapter sequences, an enzymatic cleanup of PCR products, a second PCR for normalization and incorporation of dual indexing tags (Nate's plates), and bead size selection. Libraries were sequenced on an Illumina MiSeq with a 500 cycle kit in paired-end mode.

Detailed library preparation protocols for both enrichment strategies are provided in Supplemental Text 3 and 4.

#### 1.3 Sequence analysis

We used the SeekDeep pipeline (Hathaway et al. 2018) to demultiplex, filter reads, and compute read counts. First, to join and extract reads by locus, we used the extractorPairedEnd function with default filtering and quality parameters and additional primer specifications (minOverlap: 6, primercoverage: 0.7, primerWithinStart: 20). Next, the qluster function with default parameters for Illumina data was used to create haplotypes with relative abundances. Final filtering was done using the processsClusters function with parameters that allow for a few low-quality mismatches and no indels, recommended by the developer for Illumina data (parameters: strictErrors, illumina, fracCutOff 0.02).

To evaluate the different laboratory protocols, the percentage of on-target reads was calculated by dividing the total number of reads that were matched to panel loci by SeekDeep for each sample by the total number of reads for that sample in the raw data.

### 2 Results

Among the various DNA preservation, extraction, and enrichment methods tested, the results point to two viable strategies, depending on parasite density. From the DBS samples, SWGA more consistently produced high percentages of on-target reads than the other enrichment methods (Figure 1A), but targeted pre-amplification led to more consistent amplification of each marker (Figure 2A). The Chelex- and kit-based extraction methods were relatively comparable in terms of on-target reads (Figure 1A) and marker amplification (Figure 2A). High percentages of on-target reads were obtained from whole blood samples for both SWGA and targeted pre-amplification (Figure 1B), but many loci did not amplify well from these samples, regardless of enrichment method (Figure 2B). From these results, it can be concluded that with this panel whole blood samples are not worth the additional effort and cost, and there is little difference in performance between Chelex-based and kit-based extraction. However, there are tradeoffs between SWGA and targeted pre-amplification, and it is not apparent which is best.

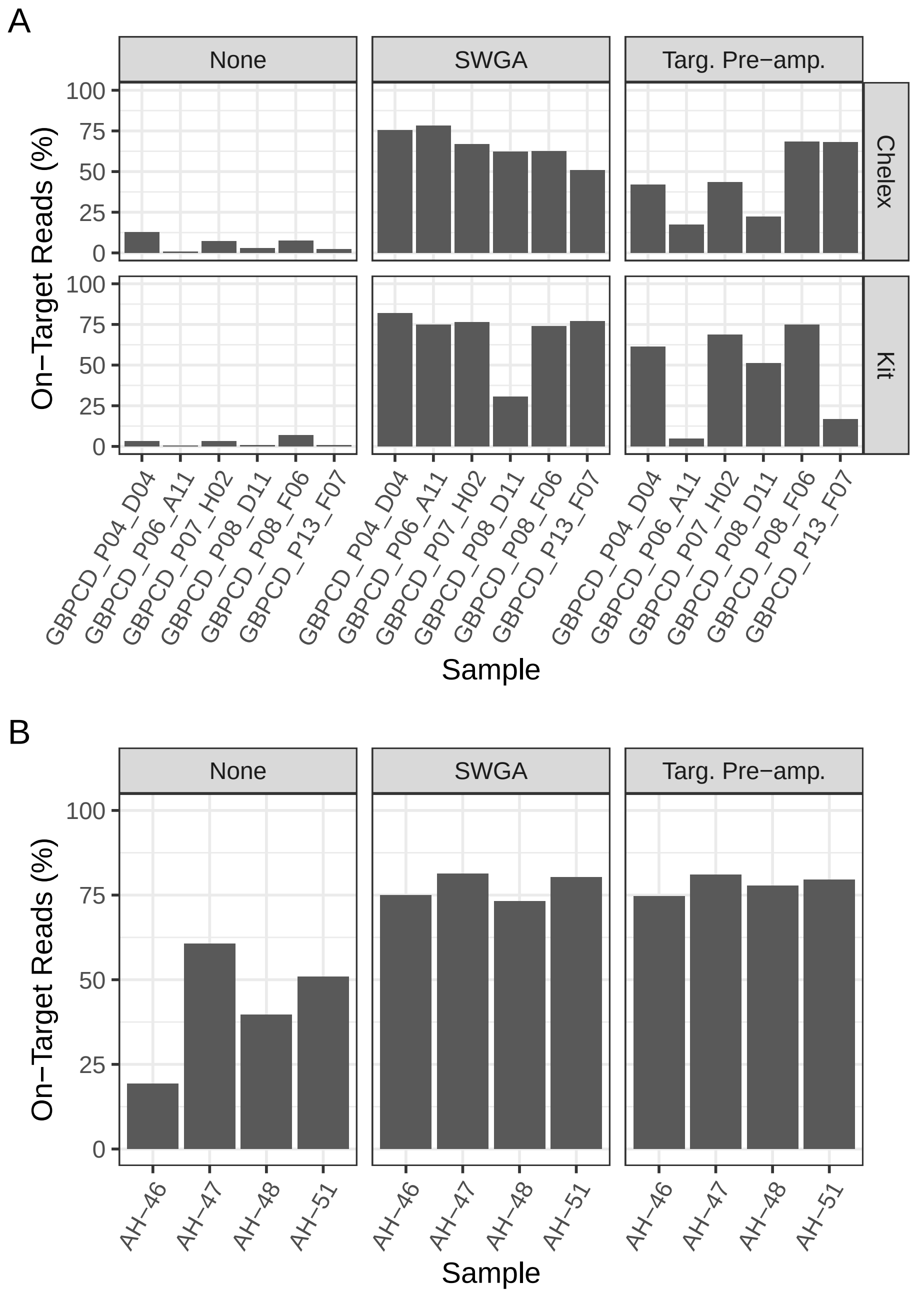


Figure 1: Bar plots indicating the percentage of on-target reads for the DBS samples (A) and the whole blood samples (B). There is one plot for each DNA enrichment method and, in the case of the DBS samples, the extraction method that was tested.

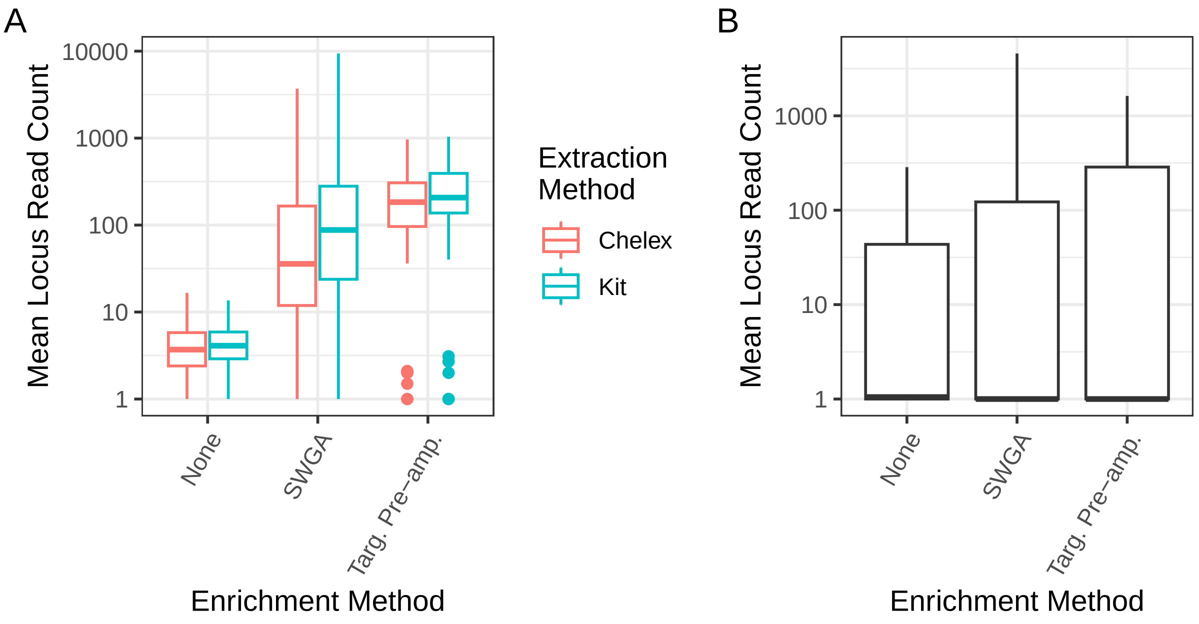


Figure 2: Box plots (with Tukey-style whiskers that extend to a maximum of 1.5 * IQR) showing the mean read count across all samples for each locus, separated according to DNA enrichment and extraction method. The DBS samples are shown in (A) and the whole blood samples in (B). In both cases, the y-axis is $log_{10}$ transformed.

The picture becomes more clear when the relationship with parasite density is considered (Figure 3). Targeted pre-amplification performs much better in DBS samples with a high parasite load, whereas the performance of SWGA is consistently high. Therefore, SWGA was selected for the final protocol, though it is noted that with high parasitemia targeted pre-amplification is also a viable option, and may in fact perform slightly better.

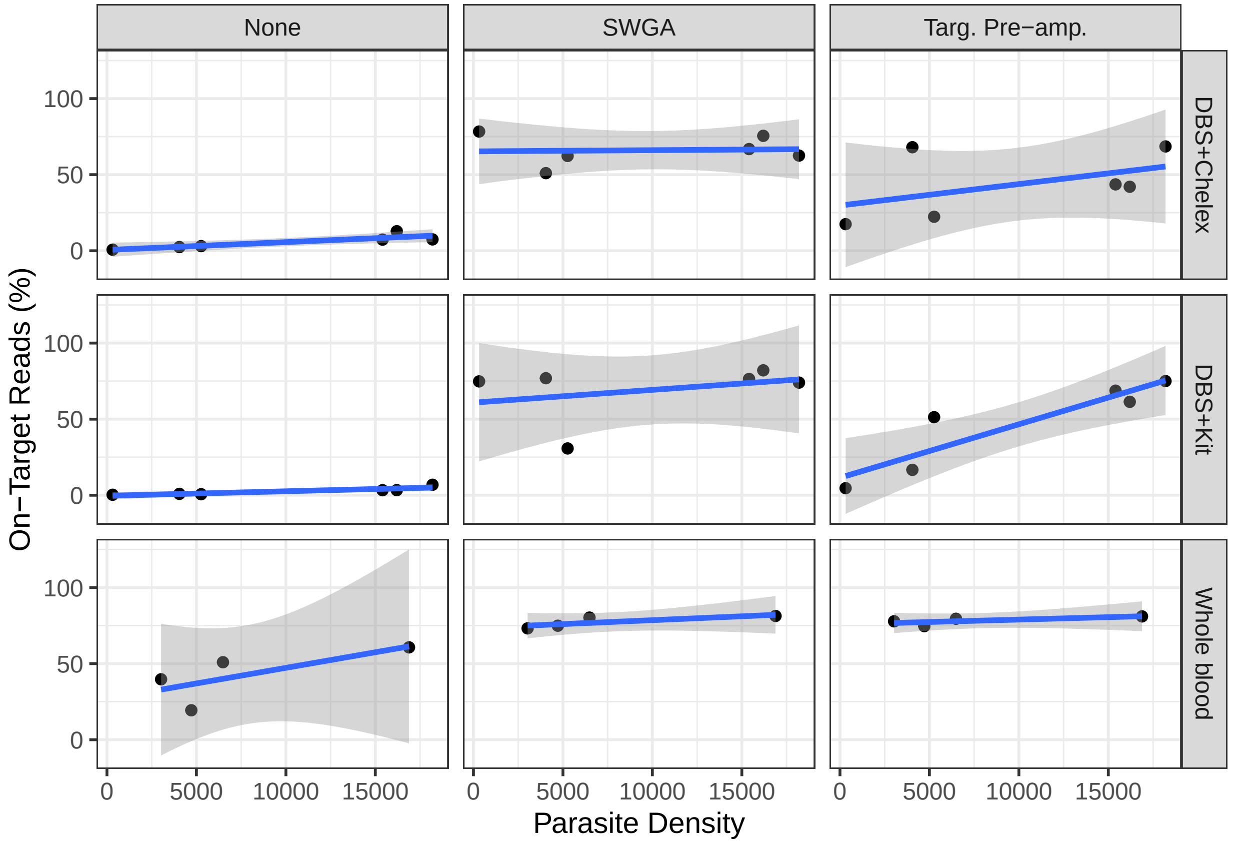


Figure 3: These scatterplots show the relationship between the percentage of on-target reads and parasite density, for each combination of enrichment protocol (none, SWGA, and targeted pre-amplification) and DNA preservation/extraction method (DBS+Chelex, DBS+Kit, and whole blood). In each case, a linear regression between parasite density and the percentage of on-target reads is displayed, with confidence intervals, to aid interpretation.
