## Supplemental Text 3 for "PvGAP: Development of a globally-applicable, highly-multiplexed microhaplotype amplicon panel for *Plasmodium vivax*"

***P. vivax* microhaplotype library preparation protocol using sWGA for enrichment** **(2026-03-05)**

**Step 1: Parasite DNA enrichment and preparation**

sWGA protocols adapted from:

- <https://doi.org/10.1186/s12936-016-1641-7>
- <https://doi.org/10.1128/mBio.02257-16>

**Step 1.1: Selective whole-genome amplification (sWGA)**

**PCR recipe:**

| Component | Concentration in 50 μL reaction | Volume (μL) |
| --- | --- | --- |
| Input gDNA | 30-70 ng (min. 10 ng) | 22 |
| sWGA primers (at 200 µM) | 3.5 µM of each primer | 0.875 each (10.5 total) |
| Phi29 DNA polymerase (NEB) | 30 units | 3 |
| 10X phi29 buffer (NEB) | 1x | 5 |
| 10 mM dNTPs | 1.8 mM | 9 |
| 100X Albumin (NEB) | 1% | 0.5 |
| NF dH20 |  | 0 |
| Total volume |  | **50** |

**PCR cycling condition:**

| Temp. (°C) | Time |
| --- | --- |
| 35 | 10:00 |
| 34 | 15:00 |
| 33 | 20:00 |
| 32 | 25:00 |
| 31 | 30:00 |
| 30 | 16:00:00 |
| 65 | 10:00 |
| 4 | Hold |

**Primers:**

Pvset1920 (* indicates phosphorothioate bonds, 5’ to 3’)

1. AACGAAGC*G*A
2. ACGAAGCG*A*A
3. ACGACGA*A*G
4. ACGCGCA*A*C
5. CAACGCG*G*T
6. GACGAAA*C*G
7. GCGAAAAA*G*G
8. GCGAAGC*G*A
9. GCGGAAC*G*A
10. GCGTCGA*A*G
11. GGTTAGCG*G*C
12. AACGAAT*C*G

**Step 1.2: Bead cleanup of sWGA products:**

Equilibrate beads (AmpureXP, CleanNGS, or Takara Nucleomag) to RT (>30 min)

Prepare fresh 80% EtOH

***NOTE: when discarding supernatant in the following steps, visually inspect pipette tip to ensure you are not discarding magnetic beads!***

a. Transfer 20µL of sWGA products to a new plate
b. Add 20µl of RT beads to sWGA product (for a 1:1 PCR product to beads ratio); pipette-mix 10-15x, quick centrifuge, and incubate for 5 min at RT.
c. Move plate to magnetic rack and let sit for 3 min at RT until supernatant is completely clear
d. Remove supernatant and discard while being careful not to discard any beads
e. Wash: while on magnetic rack, add 200 μL of 80% EtOH (freshly prepared); incubate for 30 s and discard.
f. Repeat wash step (*step e*)
g. After the second wash, spin down briefly, return to magnet, and use a P10 to remove any residual droplets.
h. Air-dry beads 2-5 min on the magnet until the pellet looks matte but not cracked. (Over-drying can reduce yield and make beads harder to elute)
i. Remove plate from magnetic rack; resuspend beads into 20 µL (or starting volume) of TE buffer; pipette mix 10-15X, quick centrifuge, and incubate for 5 min at RT
j. Move plate to magnetic rack and let sit for 3 min at RT until eluate clears
k. Collect 20 µl of supernatant in a new plate

**Step 2: GT-seq PCR**

GT-seq protocol adapted from:

- <https://doi.org/10.1111/1755-0998.12357>
- <https://doi.org/10.1111/1755-0998.13622>

**Step 2.1: PCR 1**

Thaw components at RT. Mix well and centrifuge briefly.

To make primer pool for PCR1: Add 2µL of each primer (suspended at 200 uM) into total volume of 1600 µL with TE buffer to get a 0.25 µM final concentration in the pool of primers.

**PCR recipe:**

| Component | Vol. (uL)^ | Vol. (uL)± |
| --- | --- | --- |
| Qiagen Plus MM (2X) | 5.0 | 7.5 |
| 0.25 uM primer pool | 1.5 | 1.5 |
| sWGA product | 3.0 | 6.0 |
| NF dH20 | 0.5 | 0.5 |
| Total Volume | 10.0 | 15.0 |

**^** used when parasite density >= 100 copies per uL

± used when parasite density < 100 copies per uL

**PCR cycling condition:**

| Step | Temp. (°C) | Time | Cycles^ | Cycles± |
| --- | --- | --- | --- | --- |
| Hot Start | **95** | 15:00 | 1 | 1 |
| Denaturation | **95** | 0:30 |  |  |
| Annealing | **57*** | 0:30 | 5 | 5 |
| Extension | **72** | 2:00 |  |  |
| Denaturation | **95** | 0:30 |  |  |
| Annealing | **65** | 0:30 | 15 | 18 |
| Extension | **72** | 0:30 |  |  |
| Hold | **10** | Hold | 1 | 1 |

* Slow cool: 5% ramp rate (~ 0.1-0.3 deg/s)

**^** used when parasite density >= 100 copies per uL

± used when parasite density < 100 copies per uL

*Optional: confirm successful PCR by performing gel electrophoresis before proceeding to Step 2. Use 3 µL of each PCR product.*

**Step 2.2: Remove remaining primers and primer dimers by bead cleanup**

Equilibrate beads (AmpureXP, CleanNGS, or Takara Nucleomag) to RT (>30 min)

Prepare fresh 80% EtOH

***NOTE: when discarding supernatant in the following steps, visually inspect pipette tip to ensure you are not discarding magnetic beads!***

a. You should have 7 µL of PCR product remaining. Add 13 µL of NF dH20 to each well to make bead cleanup easier. Then, add 16 µL of beads for a 1:0.8 ratio; pipette-mix 10-15x, quick centrifuge, and incubate for 5 min at RT.
b. Move plate to magnetic rack and let sit for 3 min at RT until supernatant is completely clear
c. Remove supernatant and discard while being careful not to discard any beads
d. Wash: while on magnetic rack, add 200 μL of 80% EtOH (freshly prepared); incubate for 30 s and discard.
e. Repeat wash step (*step d*)
f. After the second wash, spin down briefly, return to magnet, and use a P10 to remove any residual droplets.
g. Air-dry beads 2-5 min on the magnet until the pellet looks matte but not cracked. (Over-drying can reduce yield and make beads harder to elute)
h. Remove plate from magnetic rack; resuspend beads into 50 µL (Note this is to get a 1:10 dilution of initial concentration) of TE buffer; pipette mix 10-15X, quick centrifuge, and incubate for 5 min at RT
i. Move plate to magnetic rack and let sit for 3 min at RT until eluate clears
j. Collect 50 µL of supernatant in a new plate and proceed directly to Step 3.

**Step 3: Nate’s Plates by GTseek**

A PCR-based normalization kit designed to incorporate dual indexing tags and return equal numbers of sequencing library constructs from each sample. This is a patent-pending protocol draft.

**Step 3.1: PCR 2**

Thaw Qiagen MM at RT. Mix well and centrifuge briefly.

a. Add 2µl Qiagen Plus MM to each well of Nate’s Plate
b. Transfer 2µL of product from Step 2.2 to Nate’s Plate.
c. Place Nate’s Plate in thermocycler

**PCR cycling condition:**

| Step | Temp. (°C) | Time | Cycles |
| --- | --- | --- | --- |
| Hot Start | **95** | 15:00 | 1 |
| Denaturation | **94** | 0:30 |  |
| Annealing | **57** | 0:30 | 2 |
| Extension | **72** | 2:00 |  |
| Denaturation | **94** | 0:30 | 18 |
| Extension | **72** | 0:45 |  |
| Extension | **72** | 2:00 | 1 |
| Hold | **4** | Hold | 1 |

**Step 3.2: Sample normalization - Bind**

a. Pool samples from each well in Nate’s Plate and collect in tube; label tube as POOL. If doing multiple plates, pool each plate separately.
b. Add 500 µL of Nate’s Plates Bead Buffer (2X) to POOL tube.
c. Vortex the tube of Streptavidin beads and ensure they are adequately homogenized.
d. Carefully transfer 1 µL of strep beads to POOL tube. Vortex and incubate POOL for 15 minutes at RT, inverting tube occasionally.
e. Place POOL tube on magnetic rack and incubate for 3 minutes.
f. Remove and discard supernatant, being careful not to disturb the bead pellet.

***NOTE: the bead pellet will be very small and difficult to see. Like a wisp of dirt. Ensure that you can see the bead pellet at every step.***

g. Wash: While on magnetic rack, add 1 mL of Nate’s Plates Wash buffer; incubate for 1 minute; and discard supernatant.
h. Repeat wash step (*step g*).
i. Resuspend beads in 20 µL sterile nuclease-free water; mix well

**Step 3.3: Sample normalization – Release**

a. Pipette 10µL of resuspended beads into a strip tube for thermal cycling and add the following components

| Component | Vol. (uL) |
| --- | --- |
| Qiagen Plus MM (2X) | 20.0 |
| 10X Bead Release primers | 4.0 |
| Nuclease-free water | 6.0 |
| Resuspended beads | 10.0 |
| Total Volume | 40.0 |

*Optional: The remaining 10 µL of resuspended beads can be stored at -20 if desired.*

b. Place strip tube in thermocycler

**PCR cycling condition:**

| Step | Temp. (°C) | Time | Cycles |
| --- | --- | --- | --- |
| Hot Start | **95** | 15:00 | 1 |
| Denaturation | **94** | 0:30 |  |
| Annealing | **60** | 0:30 | 6 |
| Extension | **72** | 0:30 |  |
| Extension | **72** | 2:00 | 1 |
| Hold | **4** | Hold | 1 |

**Step 3.4: Bead size selection**

Equilibrate beads (AmpureXP, CleanNGS, or Takara Nucleomag) to RT (>30 min)

Prepare fresh 80% EtOH

***NOTE: when discarding supernatant in the following steps, visually inspect pipette tip to ensure you are not discarding magnetic beads!***

a. Transfer PCR product to a new tube and place on magnetic rack. Let sit for 3 minutes
b. Transfer 30 µL cleared supernatant to new tube. Add 21 uL beads for a 1:0.7 ratio; mix well and incubate at RT for 5 minutes.
c. Move tube to magnetic rack and let sit for 3 min at RT until supernatant is completely clear
d. Remove supernatant and discard while being careful not to discard any beads
e. Wash: while on magnetic rack, add 200 μL of 80% EtOH (freshly prepared); incubate for 30 s and discard.
f. Repeat wash step (*step e*)
g. After the second wash, spin down briefly, return to magnet, and use a P10 to remove any residual droplets.
h. Air-dry beads 2-5 min on the magnet until the pellet looks matte but not cracked. (Over-drying can reduce yield and make beads harder to elute)
i. Remove plate from magnetic rack; resuspend beads into 20 µL of TE buffer; pipette mix 10-15X, quick centrifuge, and incubate for 5 min at RT
j. Move plate to magnetic rack and let sit for 3 min at RT until eluate clears
k. Collect 20 µL of supernatant in a new tube
l. Quantify and QC the library
