## Supplemental Text 4 for "PvGAP: Development of a globally-applicable, highly-multiplexed microhaplotype amplicon panel for *Plasmodium vivax*"

***P. vivax* microhaplotype library preparation protocol using nested PCR for enrichment (2026-03-05)**

*Note that this protocol has NOT been optimized for low parasite density samples (<2500 parasite copies per µL)*

**Step 1: Parasite DNA enrichment and preparation**

**Step 1.1: Nested PCR**

Thaw components at RT. Mix well and centrifuge briefly.

To make primer pool for Nested PCR: Add 2µL of each primer (suspended at 200 uM) into total volume of 1600 µL with TE buffer to get a 0.25 µM final concentration in the pool of primers.

**PCR recipe:**

| Component | Vol. (uL) |
| --- | --- |
| Qiagen Plus MM (2X) | 5.0 |
| 0.25 uM primer pool | 1.5 |
| Genomic DNA | 3.0 |
| NF dH20 | 0.5 |
| Total Volume | 10.0 |

**PCR cycling condition:**

| Step | Temp. (°C) | Time | Cycles |
| --- | --- | --- | --- |
| Hot Start | **95** | 15:00 | 1 |
| Denaturation | **95** | 0:30 |  |
| Annealing | **57*** | 0:30 | 5 |
| Extension | **72** | 2:00 |  |
| Denaturation | **95** | 0:30 |  |
| Annealing | **65** | 0:30 | 15 |
| Extension | **72** | 0:30 |  |
| Hold | **10** | Hold | 1 |

* Slow cool: 5% ramp rate (~ 0.1-0.3 deg/s)

**Step 1.2: Dilution of nested PCR product (no cleanup):**

a. You should have 10 µL of PCR product. Add 20 µL of NF dH20 to create a 1:3 dilution. Pipette-mix 10-15x and quick centrifuge. You will use this diluted PCR product in the GT-seq PCR reaction.
b. Confirm successful PCR by electrophoresis before proceeding to Step 2

**Step 2: GT-seq PCR**

GT-seq protocol adapted from:

- <https://doi.org/10.1111/1755-0998.12357>
- <https://doi.org/10.1111/1755-0998.13622>

**Step 2.1: PCR 1**

Thaw components at RT. Mix well and centrifuge briefly.

To make primer pool for PCR1: Add 2µL of each primer (suspended at 200 uM) into total volume of 1600 µL with TE buffer to get a 0.25 µM final concentration in the pool of primers.

**PCR recipe:**

| Component | Vol. (uL) |
| --- | --- |
| Qiagen Plus MM (2X) | 5.0 |
| 0.25 uM primer pool | 1.5 |
| Diluted PCR product | 3.0 |
| NF dH20 | 0.5 |
| Total Volume | 10.0 |

**PCR cycling condition:**

| Step | Temp. (°C) | Time | Cycles |
| --- | --- | --- | --- |
| Hot Start | **95** | 15:00 | 1 |
| Denaturation | **95** | 0:30 |  |
| Annealing | **57*** | 0:30 | 5 |
| Extension | **72** | 2:00 |  |
| Denaturation | **95** | 0:30 |  |
| Annealing | **65** | 0:30 | 15 |
| Extension | **72** | 0:30 |  |
| Hold | **10** | Hold | 1 |

* Slow cool: 5% ramp rate (~ 0.1-0.3 deg/s)

*Optional: Confirm successful PCR by performing gel electrophoresis before proceeding to Step 2. Use 3 µL of each PCR product.*

**Step 2.2: Remove remaining primers and primer dimers by bead cleanup**

Equilibrate beads (AmpureXP, CleanNGS, or Takara Nucleomag) to RT (>30 min)

Prepare fresh 80% EtOH

***NOTE: when discarding supernatant in the following steps, visually inspect pipette tip to ensure you are not discarding magnetic beads!***

c. You should have 7 µL of PCR product remaining. Add 13 µL of NF dH20 to each well to make bead cleanup easier. Then, add 16 µL of beads for a 1:0.8 ratio; pipette-mix 10-15x, quick centrifuge, and incubate for 5 min at RT.
d. Move plate to magnetic rack and let sit for 3 min at RT until supernatant is completely clear
e. Remove supernatant and discard while being careful not to discard any beads
f. Wash: while on magnetic rack, add 200 μL of 80% EtOH (freshly prepared); incubate for 30 s and discard.
g. Repeat wash step (*step d*)
h. After the second wash, spin down briefly, return to magnet, and use a P10 to remove any residual droplets.
i. Air-dry beads 2-5 min on the magnet until the pellet looks matte but not cracked. (Over-drying can reduce yield and make beads harder to elute)
j. Remove plate from magnetic rack; resuspend beads into 50 µL (Note this is to get a 1:10 dilution of initial concentration) of TE buffer; pipette mix 10-15X, quick centrifuge, and incubate for 5 min at RT
k. Move plate to magnetic rack and let sit for 3 min at RT until eluate clears
l. Collect 50 µL of supernatant in a new plate and proceed directly to Step 3.
