## Supplemental Text 5 for "PvGAP: Development of a globally-applicable, highly-multiplexed microhaplotype amplicon panel for *Plasmodium vivax*"

Supplemental Text 5: Simulation-based evaluation of multiplexing capacity across sequencing kits

Alfred Hubbard Edwin Solares Lauren Bradley Brook Jeang
Delenasaw Yewhalaw Daniel Janies Eugenia Lo Guiyun Yan
Elizabeth Hemming-Schroeder

This document describes a simulation-based evaluation of the number of samples that could be multiplexed per sequencing run while maintaining adequate locus coverage across samples. The analysis uses empirical locus-level read count data from the PvGAP dataset to model unevenness in read allocation among samples and loci under different sequencing kit capacities.

#### 1 Methods

We used empirical locus-level on-target read counts from the PvGAP dataset as the basis for a resampling framework to estimate multiplexing performance across sequencing platforms. For each observed sample, we first calculated the total number of on-target reads and then converted locus-specific read counts into within-sample locus proportions. These locus proportions represent the relative distribution of reads across loci for a given sample.

To capture sample-to-sample unevenness in sequencing yield, we calculated a relative sample efficiency term for each observed sample as its total on-target read count divided by the mean total on-target read count across all samples in the empirical dataset. These efficiency terms were then resampled with replacement to generate realistic variation in how total usable reads might be distributed across multiplexed samples.

For each candidate multiplex level and sequencing kit, total reads per run were first reduced by 30% to account for reads unavailable for target genotyping, including 10% PhiX and 20% off-target amplification. The remaining reads were treated as usable on-target reads. Within each simulation replicate, sample efficiency terms were drawn with replacement to allocate the usable reads among samples, and empirical locus profiles were drawn with replacement to represent locus-to-locus unevenness within samples. Expected read counts for each locus were then obtained by multiplying the resampled locus proportions by the expected number of reads allocated to each sample.

For each simulated sample, we calculated the minimum locus read count, the number of loci meeting a minimum threshold of 10 reads, and the proportion of loci meeting that threshold. A sample was considered to pass if at least 75% of loci met the 10-read threshold. For each multiplexing level and sequencing kit, we performed 100 simulation replicates. For each metric, we summarized results across simulation replicates using the median and interquartile range (IQR). Summary metrics included the mean, median, minimum, and maximum expected reads per sample; the proportion of samples passing the locus coverage criterion; the median proportion of loci passing; the median number of loci passing; and the median minimum locus read count.

We evaluated four sequencing kit configurations: MiSeq v2 300-cycle, NextSeq 2000 P1 300-cycle, NextSeq 2000 P2 300-cycle, and NextSeq 2000 P3 300-cycle. Rather than applying a single shared sample-count grid across kits, candidate multiplexing levels were evaluated using kit-specific search ranges chosen to reflect the expected throughput of each platform. The evaluated ranges were 96–184 samples for MiSeq v2, 768–1056 samples for NextSeq 2000 P1, 3168–3936 samples for NextSeq 2000 P2, and 9408–11424 samples for NextSeq 2000 P3. To reduce unnecessary computation, evaluation within each kit stopped once the median proportion of loci passing dropped below the required threshold of 75%. The recommended multiplexing level for each kit was defined as the largest number of samples for which at least 80% of simulated samples passed the locus coverage criterion.

For the cost summary, per-sample sequencing cost was calculated by dividing the kit price by the recommended number of samples per run, and combined library preparation plus sequencing cost was calculated by adding $15 per sample in library preparation costs. Kit prices were based on internal pricing provided by the Colorado State University Next-Generation Sequencing Core.

### 2 Results

Simulation-based resampling of empirical PvGAP locus coverage profiles was used to evaluate how multiplexing level influences the proportion of samples achieving adequate locus coverage across sequencing platforms. As expected, increasing the number of multiplexed samples per run reduced the proportion of samples meeting the locus coverage criterion, reflecting declining per-sample sequencing depth as reads are distributed across larger sample pools (Figure 1).

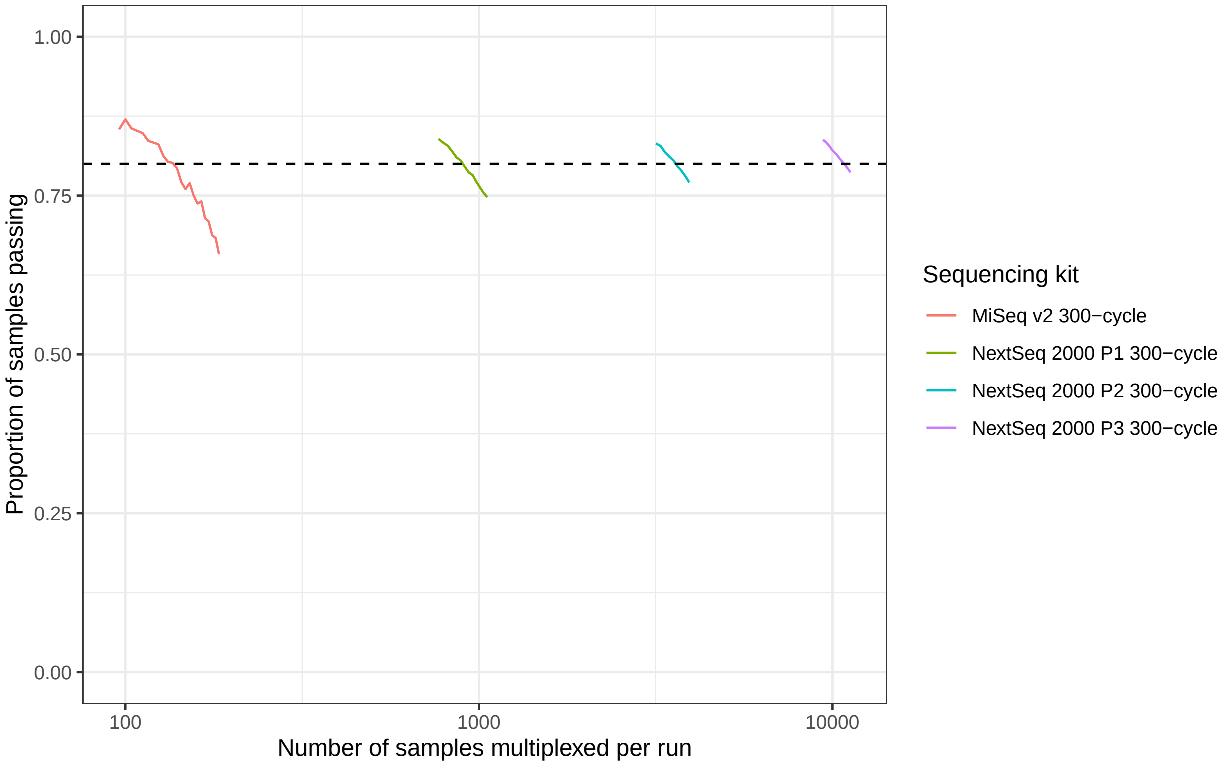


Figure 1: Estimated proportion of simulated samples passing the locus coverage criterion across candidate multiplexing levels for each sequencing kit. The horizontal dashed line indicates the threshold used to define the recommended maximum multiplexing level (80% of samples passing).

Across all evaluated sequencing platforms, acceptable multiplexing levels could be identified where at least 80% of simulated samples had coverage of at least 75% of loci with a minimum of 10 reads. Recommended multiplexing levels and associated performance metrics are summarized in Table 1. These values represent the largest number of samples per run for which the defined locus coverage criterion was satisfied based on the median simulation results.

Estimated per-sample sequencing costs were calculated by dividing reagent kit prices by the recommended number of samples per run, and combined library preparation plus sequencing costs were calculated by adding $15 per sample in library preparation costs. Estimated costs for each platform are summarized in Table 1.

Overall, the simulation results indicate that high levels of multiplexing enable cost-effective implementation of the PvGAP panel across sequencing platforms while maintaining acceptable locus coverage.

Table 1: Recommended maximum multiplexing level for each sequencing kit. Values for samples passing and loci passing are reported as median (Q1–Q3) across 100 simulation replicates.

| Sequencing kit | Samples per run | Samples passing (%) | Loci passing (%) | Sequencing cost per sample ($) | Library prep + sequencing per sample ($) |
|---|---|---|---|---|---|
| MiSeq v2 300-cycle | 136 | 80.1 (75.7–83.3) | 85.9 (84.7–87.1) | 13.24 | 28.24 |
| NextSeq 2000 P1 300-cycle | 888 | 80.5 (79.3–82.0) | 85.9 (85.9–87.1) | 2.47 | 17.47 |
| NextSeq 2000 P2 300-cycle | 3552 | 80.5 (79.8–81.3) | 85.9 (85.9–85.9) | 1.04 | 16.04 |
| NextSeq 2000 P3 300-cycle | 10696 | 80.2 (79.9–80.6) | 85.9 (85.9–85.9) | 0.51 | 15.51 |
